# Relationship between high rates of intestinal parasitic infections and knowledge, attitudes, and practices of Ndelele Health District populations (East Region, Cameroon): a cross-sectional mixed approach

**DOI:** 10.1101/2021.11.29.21267024

**Authors:** Viviane Ongbassomben, Cyrille Ndo, Ericka A. Lebon, Hugues C. Nana Djeunga, Albert L. Same Ekobo, Dieudonné Adiogo

## Abstract

**Background:** Intestinal parasitic infections remain of public health concern worldwide, especially among rural and poorest populations as a consequence of precariousness, lack of sanitation, non-availability of potable water and poor hygiene conditions. The present study aimed to better understand the epidemiology of intestinal parasitic infections in rural areas of forested Cameroon.

**Methodology:** A cross-sectional survey was conducted in three Health Areas (Ndelele, Kentzou and Lolo) of the Ndelele Health District (East Region, Cameroon). Information on socio-demographic characteristics, knowledge, attitudes, and practices regarding intestinal parasitic infections were collected using a semi-structured questionnaire. Stool samples were collected and analyzed by the Kato-Katz and formalin-ether concentration techniques to complement simple direct examination.

**Principal Findings:** A total of 406 individuals belonging to three main groups (Kako or Bantu, Baka or Pygmies and Central African Republic refugees) were enrolled in the study. The overall intestinal parasitic infection rate was 74.9%, including 57.2% cases of polyparasitism. Fourteen parasite species were identified, 89.1% being intestinal protozoa and 41.8% belonging to helminths. Infections with helminths were associated with Baka (P < 0.0001). Spring water consumption was associated with hookworm infection (OR = 3.87; P = 0.008). Garbage deposited near houses was positively associated with infection with *Giardia lamblia* (OR = 3.41; P = 0.003). Polyparasitism was positively linked to washing hand without soap before meal (OR= 11.64; p= 0.002).

**Conclusion/Significance:** Intestinal parasitic infections exhibited high rates in the Ndelele Health District, especially among indigenous and hard-to-reach populations (Pygmies). Hygiene measures appear as the main drivers sustaining transmission, and targeted strategies should be developed to efficiently fight against these debilitating diseases.

**Author summary:** Intestinal parasitic infections such as soil transmitted helminthiasis and schistosomiasis, remain a public health concern in Cameroon. To better understand the epidemiology of these infections in hard-to-reach populations in rural areas, a cross-sectional study was carried out in three health areas of Ndelele health District in the Eastern-Cameroon. The rate of intestinal parasitic infections was high (74.9%) especially in indigenous populations (Baka-Pygmies) which exhibited high prevalence of soil transmitted helminth infections (83.3%). The study confirmed that the lack of sanitation and poor hygiene largely contribute to the endemicity of intestinal parasitic infections, particularly among indigenous populations. Targeted control strategies seem mandatory to reach these populations and offer them appropriate care to interrupt or reduce the transmission of these diseases.

## Introduction

Intestinal parasitosis are infections caused by intestinal helminths (soil transmitted helminth or *Schistosoma spp* infections) and protozoa. They are widespread in developing countries, particularly in the tropics. Absence of sanitation networks, lack of drinking water, poor public and private hygiene standards, and insufficient health education exacerbate infection with eggs, larvae and cysts through contact with contaminated soil, food or water [1-2]. More than 1.5 billion people representing 24% of the world’s population are infected with soil-transmitted helminth infections worldwide [3]. Estimates show that at least 290.8 million people required preventive treatment for schistosomiasis in 2018, out of which more than 97.2 million have received appropriate treatment [4]. The main species of soil-transmitted helminths that infect humans are roundworm (*Ascaris lumbricoides*), whipworm (*Trichuris trichiura*) and hookworms (*Necator americanus* and *Ancylostoma duodenale*) [5]. The most important intestinal protozoan infections are amoebiasis (*Entamoeba histolytica*), giardiasis (*Giardia lamblia*), cryptosporidiosis (*Cryptosporidium sp*), cyclosporiasis (*Cyclospora cayetanensis*) and isosporiasis (*Isospora belli*) [6]. Children of pre-school and school ages are the most affected by intestinal parasitosis [7-8]. Over 270 million preschool-age children live in areas where these diseases are intensively transmitted [9]. Helminths negatively influence the nutritional status of children and adolescents as they inhibit absorption of nutrients, leading to stunted growth [10]. *Cryptosporidium spp*. is most common in children with diarrhea, followed by *G. lamblia* and *E. histolytica* [11-12]. The strategy deployed by the World Health Organization (WHO) to control soil-transmitted helminth infections is periodic mass administration of Albendazole or Mebendazole to at-risk populations (pre-school age children, school age children, women of reproductive age, adults in certain high-risk occupations such as tea-pickers or miners) living in endemic areas [3-13].

In Cameroon, intestinal parasitosis remains a public health concern. WHO report on the estimation of helminthic infections in Cameroon illustrates the unequal distribution of helminthiasis across the country. Schistosomiasis is highly endemic in the Northern part of the country. *Ascaris lumbricoides* and *T. trichiura* are widely distributed in the Centre and South Regions of the country, with the highest prevalence found in the South [14]. Although studies are carried out in some localities [15,16,17], updated and detailed epidemiological data on intestinal parasitic infections across the country are quite scanty. This is due not only to limited funding but also to the difficulty to access certain zones due to very poor road network. This is especially the case of the East Region of Cameroon which is very landlocked rendering implementation of epidemiological surveys difficult. As consequence, there is lack of data on the epidemiology of several tropical diseases in this region, and people usually feel like they are underlooked. This study was therefore conducted to provide unprecedented detailed data on the epidemiology of intestinal parasitic infections in a hard-to-reach Health District of the East Region of Cameroon.

## Methods

### Ethical considerations

An ethical clearance (N°1750 CEI-UDo/04/2019/M) was granted by the Institutional Review Board (IRB) of the University of Douala. Thereafter, administrative authorizations were delivered by the East Regional Delegate for Public Health, the Ndelele District Medical Officer and the Heads of the Ndelele and Kentzou Sub-Divisions. Prior to enrolment, participants were briefed about the study objectives and design. All adult volunteers who agreed to participate signed a written informed consent form, while formal consent was obtained from the parent or legal guardian for children under the age of 18. A unique identifier was used for each participant for anonymity purpose.

### Study area and population

The present study was conducted in the Ndelele Health District located in the Kadey Division, East Region, Cameroon. This Health District is bordered to the North by the Batouri Health District, to the South by the Yokadouma Health District, to the West by the Mbang Health District and to the East by the Bombette and the Kadey river which border the Central African Republic. In the study area, prevails a Guinean type tropical climate divided into four seasons: a short rainy season extending from March to June, a short dry season from July to August, a long rainy season from September to November, and a long dry season from December to March. Vegetation is mainly made up of dense forest in the Ndelele Health area, and wooded savannah sprinkled with forest galleries in Kentzou and Lolo Health areas.

According to the 2019 census, the population of the Ndelele Health District, including Central African refugees, was 63,319 inhabitants. This population is organized in several ethnic groups, of which the most important are the Kako (Bantu), which represents a large majority of the population, the Baka (Pygmies), an indigenous minority groups mostly settled in the forest, and an important community of Central African Bororo refugees. Economic activities of the area are mainly based on agriculture, mining, animal husbandry, logging, hunting, and trading are also practiced in the Ndelele Health District [18-19].

### Study design

A household-based cross-sectional survey was carried out in the Ndelele Health District to investigate the prevalence, intensity of infection and risk factors associated with intestinal parasitic infections. All volunteers, whatever their age and gender, living in the Ndelele, Kentzou and Lolo Health areas of the Ndelele Health District, were eligible to this study. A semi-structured questionnaire (translated in local language by the community health workers when necessary) was administered to capture their socio-demographic characteristics and potential factors associated with transmission of intestinal parasitic infections (WaSH indicators, gender, age groups, ethics groups and health areas). One part of the questionnaire was only administered to participants over 14 years old to know their knowledge about the disease. The WaSH part (hand washing, water supply, defecation place, barefoot walking) of children under 7 years old was answered by parents. All participants with seven years or more were responding to the WaSH part of the questionnaire

### Sample collection and processing

After the interview, a labelled sterile 60mL plastic screw-cap vial was provided to the participants (or their parents or legal guardian when they were minors), and stool samples were collected the next day in the morning, between 7 and 8 am. Sample were first analyzed by direct observation of fresh stool in saline (NaCl 0.9%) to search for worm eggs, larvae, protozoan trophozoïtes and cysts. When cyst were present, an iodine wet mount was performed for further their identification by staining their glycogen and nuclei [20]. Two concentration techniques were subsequently performed for the diagnosis of intestinal helminths [21]. Firstly and using the Kato Katz technique thick smears were prepared on microscope slides using standard 41.7 mg templates. Slides were first examined under microscope (magnification x100 or x400) within 30 minutes for the detection of hookworm eggs, then 24 hours later for the identification of other helminth eggs. When present, eggs were identified and counted, and the results expressed as number of eggs per gram of stool (epg). Secondly, stool samples were processed by Formalin-ether concentration technique. Indeed, a suspension of 1g of stool mixed with 10 mL of 10% formalin was prepared and strained through sieve in centrifugation tube; formalin was added in centrifugation tube to adjust the total volume to 10 mL. After adding 3 mL of ether to the centrifugation tube, the tube was vortexed and then centrifuged for 3 min at 500g. From the resulting four layers, the three top layers were discarded, and the resulting sediment was examined using bright field microscopy for detection of helminths eggs and intestinal protozoa cysts [22].

### Statistical analysis

Data were entered in a purpose-built Microsoft Office Excel sheet and were exported to Statistical Package for the Social Sciences (SPSS) version 23 software (SPSS Inc, Chicago, IL, USA) for analyses. Infection rates were expressed as the number of people infected by at least one parasite in the study population. For STH specifically, intensities of infection were classified according to WHO guidelines, as follows: light, moderate and heavy infections when the epg was 1–1,999, 2,000–3,999 and ≥4,000 for *hookworm*; 1– 4,999, 5,000–49,999 and ≥50,000 for *A. lumbricoides*; and 1–999, 1,000–9,999 and ≥10,000 for *T. trichiura* [23]. Chi-square test was used to assess the difference in infection rates and proportions of intensities of infection categories according to age groups, genders, Heath Areas, ethnic groups and WASH indicators (use of latrine, open defecation and water sources). Logistic regression model was used to investigate the association between parasite infections and socio-demographic factors (gender, age, ethnic group and Health Area) as well as WASH indicators adjusted with age, gender, ethnicity, and health area. Threshold for significance was set at 0.05.

## Results

### Socio-demographics characteristics

A number of 406 participants were included in the study, (166 males and 240 females). Flighty nine (14.5%) were aged <5 years (preschool-aged children), 98 (24.1%) were aged 5-14 years (school-aged children) and 249 (61.4%) were aged >14 years (adolescents and adults). The enrollees were undereducated, with 7.6% of participants having reached high school, while most of the interviewees have never attended school (34.2%) or have attended primary school (38.4%) or Koranic schools (19.7%). The majority of participants (43.6%) lived in Ndelele Health area, while 26.6% and 29.8% were recruited in Kentzou and Lolo Health areas, respectively. Regarding the ethnic groups, most of them were Kako (47.8%) and refugees from the Central African Republic (35.5%), Baka (pygmies; 11.1%) and other ethnic groups (5.7%) being underrepresented as it is the case in the general population.

### WASH indicators

All the interviewees declared practicing hand washing before meals (100%), though mostly without soap (96.6%). Although the borehole water was the most used as drinking water (63.1%), spring water consumption was significantly higher in Ndelele Health area (50.3%) (p<0.0001). The number of households without latrine (37.1%) and the proportion of people practicing open defecation (50.3%) were significantly higher in the Ndelele Health Area. As most of Baka households were without latrines (87.5%), this ethnic group practiced open defecation more than other ethnic groups (88.9%) (<0.0001) (Table 1). Barefoot walking (76.3%) and open defecation (45.8%) were practiced mostly by pre-school aged children (p=0.0001) (Table 2).

**Table 1:**
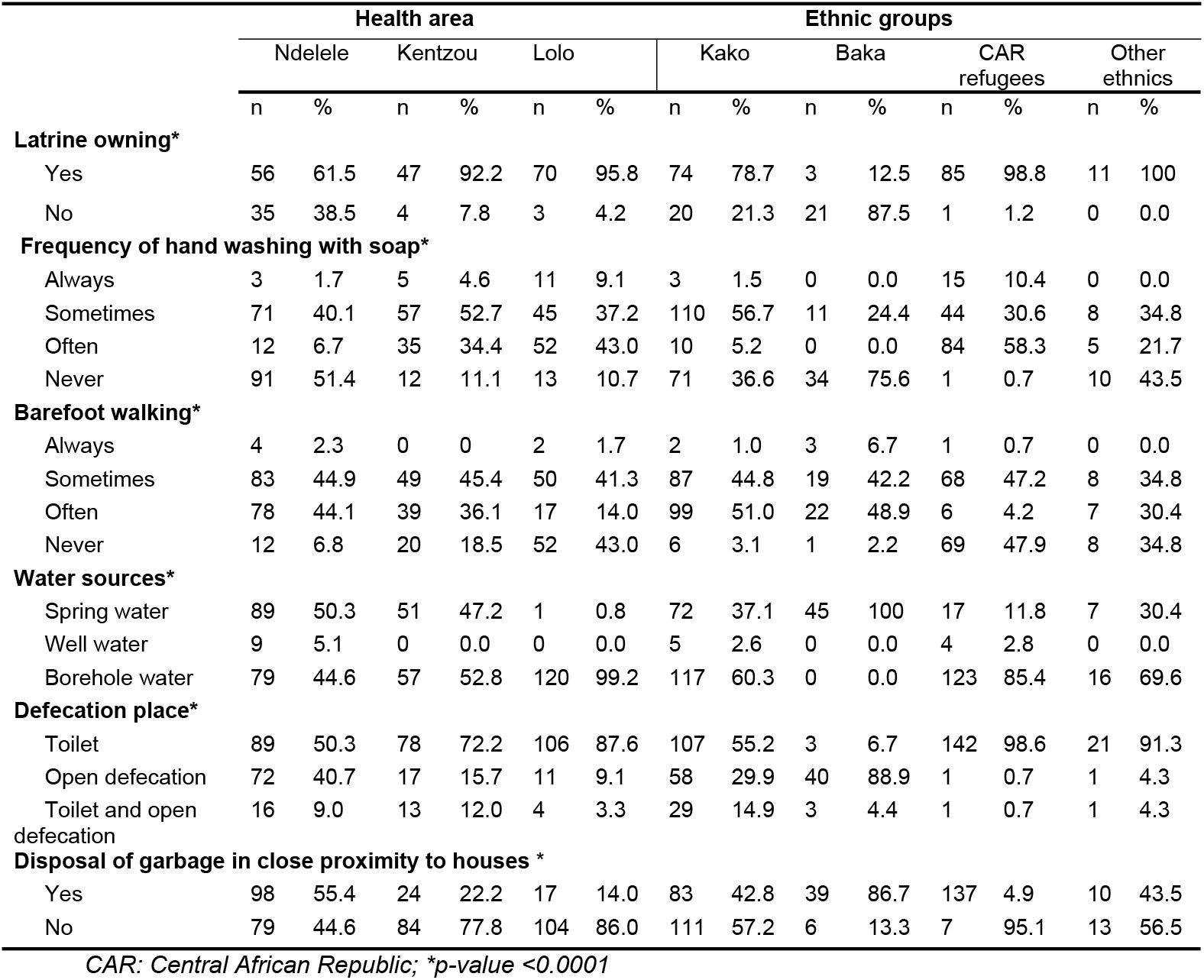
WASH indicators according to Health area and ethnic groups.

**Table 2:**
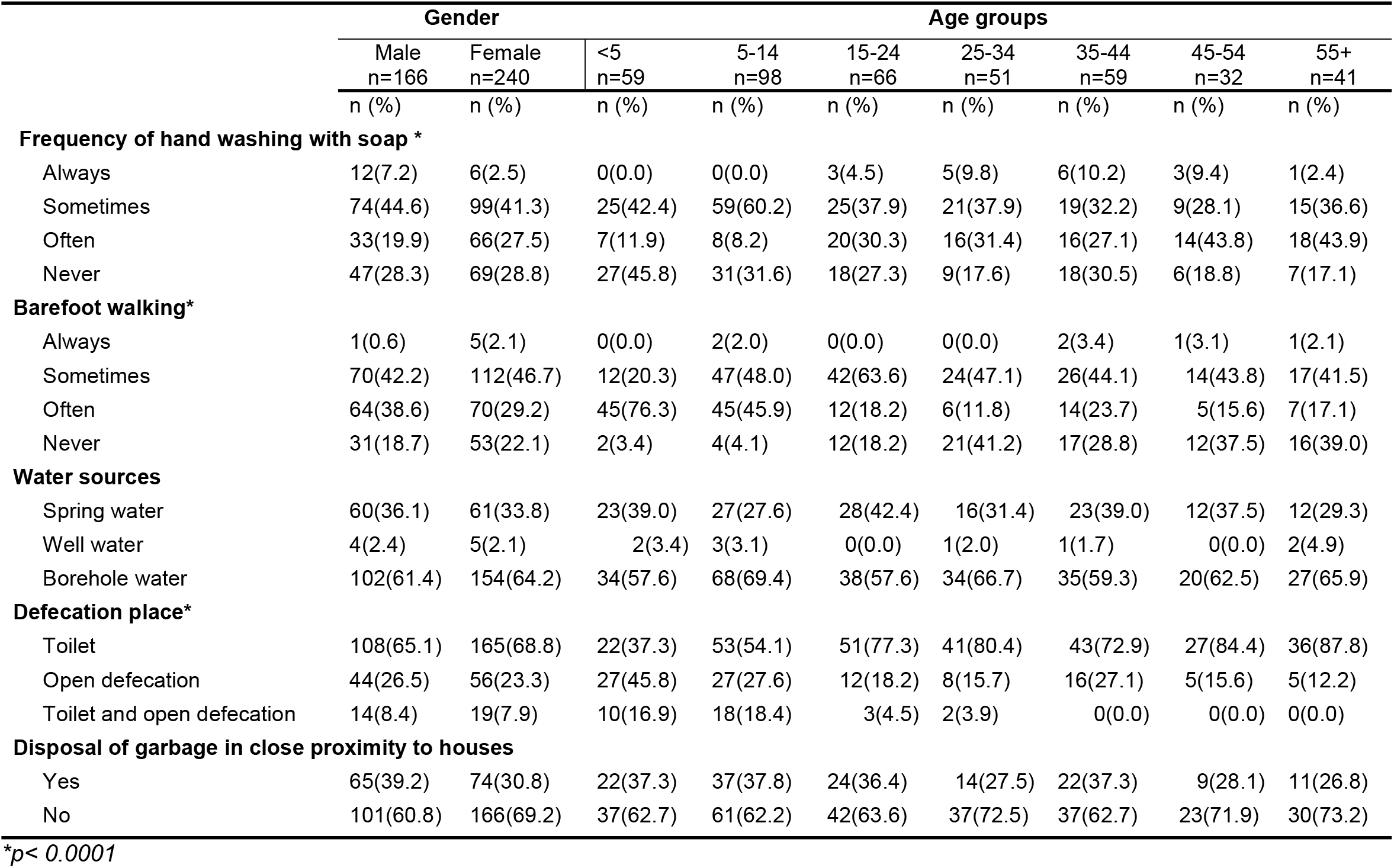
WASH indicators according to gender and age groups.

|  | Gender |  | Age groups |  |  |  |  |  |  |
| --- | --- | --- | --- | --- | --- | --- | --- | --- | --- |
|  | Male<br>n=166<br>n (%) | Female<br>n=240<br>n (%) | <5<br>n=59<br>n (%) | 5-14<br>n=98<br>n (%) | 15-24<br>n=66<br>n (%) | 25-34<br>n=51<br>n (%) | 35-44<br>n=59<br>n (%) | 45-54<br>n=32<br>n (%) | 55+<br>n=41<br>n (%) |
| **Frequency of hand washing with soap \*** |  |  |  |  |  |  |  |  |  |
| Always | 12(7.2) | 6(2.5) | 0(0.0) | 0(0.0) | 3(4.5) | 5(9.8) | 6(10.2) | 3(9.4) | 1(2.4) |
| Sometimes | 74(44.6) | 99(41.3) | 25(42.4) | 59(60.2) | 25(37.9) | 21(37.9) | 19(32.2) | 9(28.1) | 15(36.6) |
| Often | 33(19.9) | 66(27.5) | 7(11.9) | 8(8.2) | 20(30.3) | 16(31.4) | 16(27.1) | 14(43.8) | 18(43.9) |
| Never | 47(28.3) | 69(28.8) | 27(45.8) | 31(31.6) | 18(27.3) | 9(17.6) | 18(30.5) | 6(18.8) | 7(17.1) |
| **Barefoot walking\*** |  |  |  |  |  |  |  |  |  |
| Always | 1(0.6) | 5(2.1) | 0(0.0) | 2(2.0) | 0(0.0) | 0(0.0) | 2(3.4) | 1(3.1) | 1(2.1) |
| Sometimes | 70(42.2) | 112(46.7) | 12(20.3) | 47(48.0) | 42(63.6) | 24(47.1) | 26(44.1) | 14(43.8) | 17(41.5) |
| Often | 64(38.6) | 70(29.2) | 45(76.3) | 45(45.9) | 12(18.2) | 6(11.8) | 14(23.7) | 5(15.6) | 7(17.1) |
| Never | 31(18.7) | 53(22.1) | 2(3.4) | 4(4.1) | 12(18.2) | 21(41.2) | 17(28.8) | 12(37.5) | 16(39.0) |
| **Water sources** |  |  |  |  |  |  |  |  |  |
| Spring water | 60(36.1) | 61(33.8) | 23(39.0) | 27(27.6) | 28(42.4) | 16(31.4) | 23(39.0) | 12(37.5) | 12(29.3) |
| Well water | 4(2.4) | 5(2.1) | 2(3.4) | 3(3.1) | 0(0.0) | 1(2.0) | 1(1.7) | 0(0.0) | 2(4.9) |
| Borehole water | 102(61.4) | 154(64.2) | 34(57.6) | 68(69.4) | 38(57.6) | 34(66.7) | 35(59.3) | 20(62.5) | 27(65.9) |
| **Defecation place\*** |  |  |  |  |  |  |  |  |  |
| Toilet | 108(65.1) | 165(68.8) | 22(37.3) | 53(54.1) | 51(77.3) | 41(80.4) | 43(72.9) | 27(84.4) | 36(87.8) |
| Open defecation | 44(26.5) | 56(23.3) | 27(45.8) | 27(27.6) | 12(18.2) | 8(15.7) | 16(27.1) | 5(15.6) | 5(12.2) |
| Toilet and open defecation | 14(8.4) | 19(7.9) | 10(16.9) | 18(18.4) | 3(4.5) | 2(3.9) | 0(0.0) | 0(0.0) | 0(0.0) |
| **Disposal of garbage in close proximity to houses** |  |  |  |  |  |  |  |  |  |
| Yes | 65(39.2) | 74(30.8) | 22(37.3) | 37(37.8) | 24(36.4) | 14(27.5) | 22(37.3) | 9(28.1) | 11(26.8) |
| No | 101(60.8) | 166(69.2) | 37(62.7) | 61(62.2) | 42(63.6) | 37(72.5) | 37(62.7) | 23(71.9) | 30(73.2) |

\* $p < 0.0001$

### Knowledge of participants with regards with intestinal parasitic infections

Of the 249 participants interviewed, 145 (79.7%) mentioned food as the main mode of contamination of intestinal parasitic infection, with significantly higher proportion (p=0.0003) in the Kentzou Health Area (97.6%), compared to Ndelele (76.5%) and Lolo (72.2%) Health Areas. The most frequent clinical sign associated with intestinal parasitosis by the interviewees were abdominal pain (39.0%), which has been mentioned by 54.8% in Kentzou, 28.1% in Lolo and 23.5% in Ndelele. The other most known symptoms were vomiting (31.3%), and diarrhea (11.5%). The Baka ethnic group (pygmies) exhibited the worst knowledge regarding intestinal parasites.

### Participants’ attitude and practice in case of suspected infection

The majority of the enrollees did not use drugs for intestinal parasitic infections (57.1%). Of the 174 (42.8%) people using medicines for intestinal parasitic infections treatment, 89 (51.1%) were using modern medicine (Mebendazole (36.2%), Metronidazole (10.3%), and Albendazole (3.4%) whereas 54 (32.8%) used to use traditional medicine such as bark of trees or leaves of plants. The use of ‘‘drugs’’ was significantly higher (p<0.0001) in the Ndélélé Health Area (79.1%) compared to Kentzou (22.2%) and Lolo (6.6%) health Areas. Baka ethnic group (Pygmies) essentially used traditional drugs in case of suspected intestinal parasitic infection.

### Prevalence of intestinal parasitic infections

A total of 406 enrollees provided stool samples for diagnostic of intestinal parasitic infection. The overall prevalence was 74.9%, and this was significantly higher in Ndelele Health Area (85.3%) compared to the other Health Areas (p<0.0001). Baka ethnic group was significantly more infected (93.3%) compared to the other ethnic groups (p<0.0001). Fourteen parasitic species were found including nine protozoa and five helminth species (Table 3). Intestinal protozoa were found in 89.5% of infected people. The main protozoan species found were *E. coli* (50.0%), *Blastocystis hominis* (48.0%) and *E. histolytica* (26.3%). Helminths were found in 41.8% of infected people, and the main species found were *A. lumbricoides* (31.2%) and *T. trichiura* (11.2%). The distribution of parasitic species was significantly higher in the Ndelele Health Area (74.4%), followed by Kentzou (64.3%) and Lolo (50%) (Fig 1). Prevalence of helminth and protozoa infections was similar between males and females (Fig 2). School-age children were significantly more infected by *A. lumbricoides* (32.7%; p=0.045) whereas pre-school age children were more likely infected by *Strongyloides stercoralis* (13.6%; p=0.001). Regarding the ethnic groups, Baka (Pygmies) were significantly more infected by helminths such as *A. lumbricoides* (83.3%; p<0.0001), *T. trichiura* (68.9%; p<0.0001) and hookworm (26.7%; p<0.0001). Although Protozoan infections were the most common among Central African refugees (97.4%; p=0.011), prevalence were lower compare to other ethnics. For example, Kako (autochthones) were most often infected by *E. coli* and *E. histolytica* than Central African Refugees, respectively [(50.0% vs 18.1%; p<0.0001) and (25.3% vs 11.1%; p=0.012)]

**Table 3:** Parasitic infections in ethnics groups.

| Parasitic species | Ethnic groups |  |  |  |  |  |  |  |
| --- | --- | --- | --- | --- | --- | --- | --- | --- |
|  | Kako (n=194) |  | Baka (n=45) |  | Central African refugees (n=144) |  | Other ethnics (n=23) |  |
|  | Infected | % | Infected | % | Infected | % | Infected | % |
| **Helminths** |  |  |  |  |  |  |  |  |
| *Ascaris lumbricoides* | 54 | 27.8 | 31 | 68.9** | 4 | 2.8 | 6 | 26.1 |
| *Trichuris trichiura* | 21 | 10.8 | 12 | 26.7** | 0 | 0.0 | 1 | 4.3 |
| Hookworm | 15 | 7.7** | 2 | 4.4 | 1 | 0.7 | 0 | 0.0 |
| *Strongyloides stercoralis* | 10 | 5.2 | 4 | 8.9 | 0 | 0.0 | 0 | 0.0 |
| *Enterobius vermicularis* | 1 | 0.5 | 0 | 0.0 | 0 | 0.0 | 0 | 0.0 |
| **Protozoa** |  |  |  |  |  |  |  |  |
| *Entamoeba Coli* | 97 | 50.0* | 19 | 42.2 | 27 | 18.8 | 9 | 39.1 |
| *Entamoeba histolytica* | 49 | 25.3** | 10 | 22.2 | 16 | 11.1 | 5 | 21.1 |
| *Entamoeba hartmanni* | 5 | 2.6 | 4* | 8.9 | 0 | 0.0 | 0 | 0.0 |
| *Endolimax nana* | 11 | 5.7 | 1 | 2.2 | 2 | 1.4 | 0 | 0.0 |
| *Iodamoeba butshlii* | 21 | 10.8* | 2 | 4.4 | 1 | 0.7 | 0 | 0.0 |
| *Giardia lamblia* | 16 | 8.2 | 2 | 4.4 | 8 | 5.6 | 3 | 13.0 |
| *Balantidium coli* | 1 | 0.5 | 0 | 0.0 | 0 | 0.0 | 0 | 0.0 |
| *Isospora belli* | 1 | 0.5 | 0 | 0.0 | 1 | 0.7 | 0 | 0.0 |
| *Blastocystis hominis* | 80 | 41.2 | 18 | 40.0 | 42 | 29.2 | 6 | 26.1 |

\* $p<0.05$ ; \*\* $p<0.0001$

**Fig. 1.**
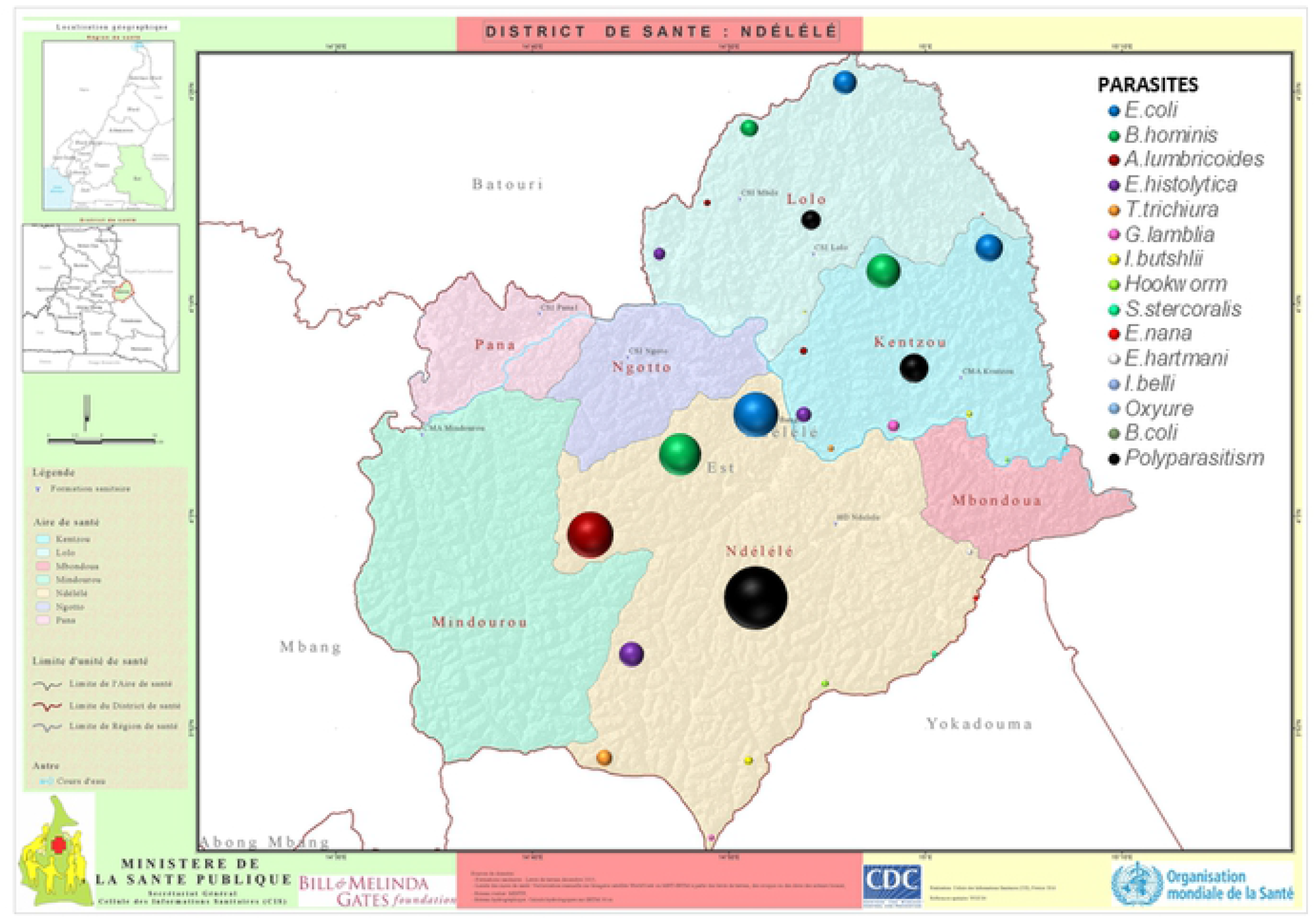
Spatial distribution of parasitic infections in Ndelele, Kentzou and Lolo health areas of Ndelele Health District.

**Fig. 2.**
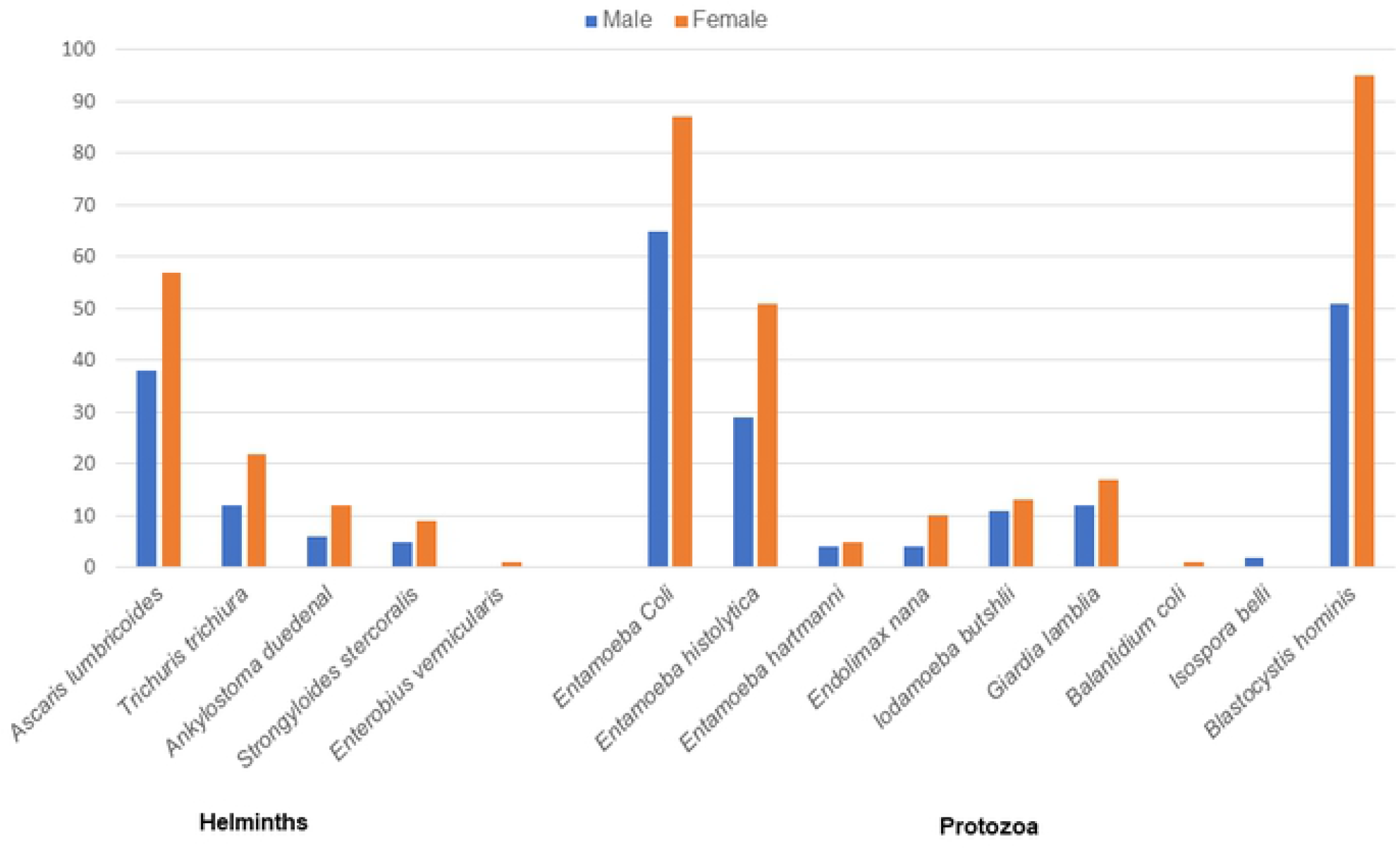
Frequency of parasitic infections according to gender.

Polyparasitism was found in 57.2% of enrollees, including infection with two (44.2%), three (37.9%) and more than four (17.8%) parasites species. Polyparasitism was similar between genders with a decreasing trend with age. It was significantly low among Central African refugees (29.9%) compared to Kako (69.5%) and Baka (66.7%) ethnic groups (p< 0.0001). Regarding ethnic groups, polyparasitism was significantly higher in Ndelele Health Area (65.6%; p =0.012).

### Intensity of helminth infections

Intensity of infection was detrmined only for helminth infections using Kato-Katz technique. This parameter varied from 96 to 76,608 epg [median: 2200; interquartile range (IQR): Q1=1000; Q3=7000] for *A. lumbricoides*, from 96 to 2,520 epg [median: 200; IQR: Q1=96; Q3=475] for *T. trichiura*, and from 96 to 3,500 epg [median: 148; IQR: Q1=96; Q3= 325] for hookworm. According to WHO classification, intensity of helminth infections was light for almost all the enrollees (77.2%) and parasite species, *A. lumbricoides* exhibiting only two cases of heavy infections in the Ndelele Health Area (Table 3).

### Association between parasitic infections, socio-demographic factors, and WASH indicators

A positive association was observed between Baka ethnic group (Pygmies) and helminth infections, especially *A. lumbricoides* (OR=4.04; P<0.0001) and *T. trichiura* (OR=2.99; p=0.007). Open defecation was positively associated with *T. trichiura* (OR=5.58; p<0.0001). Spring water consumption was positively associated with hookworm infection (OR=3.87; p=0.008) (Table 4). Infection with *G. lamblia* was positively linked to disposal of garbage near houses (OR=3.41; p= 0.003) and Kentzou Health Area (OR=3.73; p=0.002). Infection with *E. coli* was positively associated with Lolo Health Area (OR=2.61; p=0.011) and open defecation (OR=2.16; p=0.001) (Table 4).

**Table 4:**
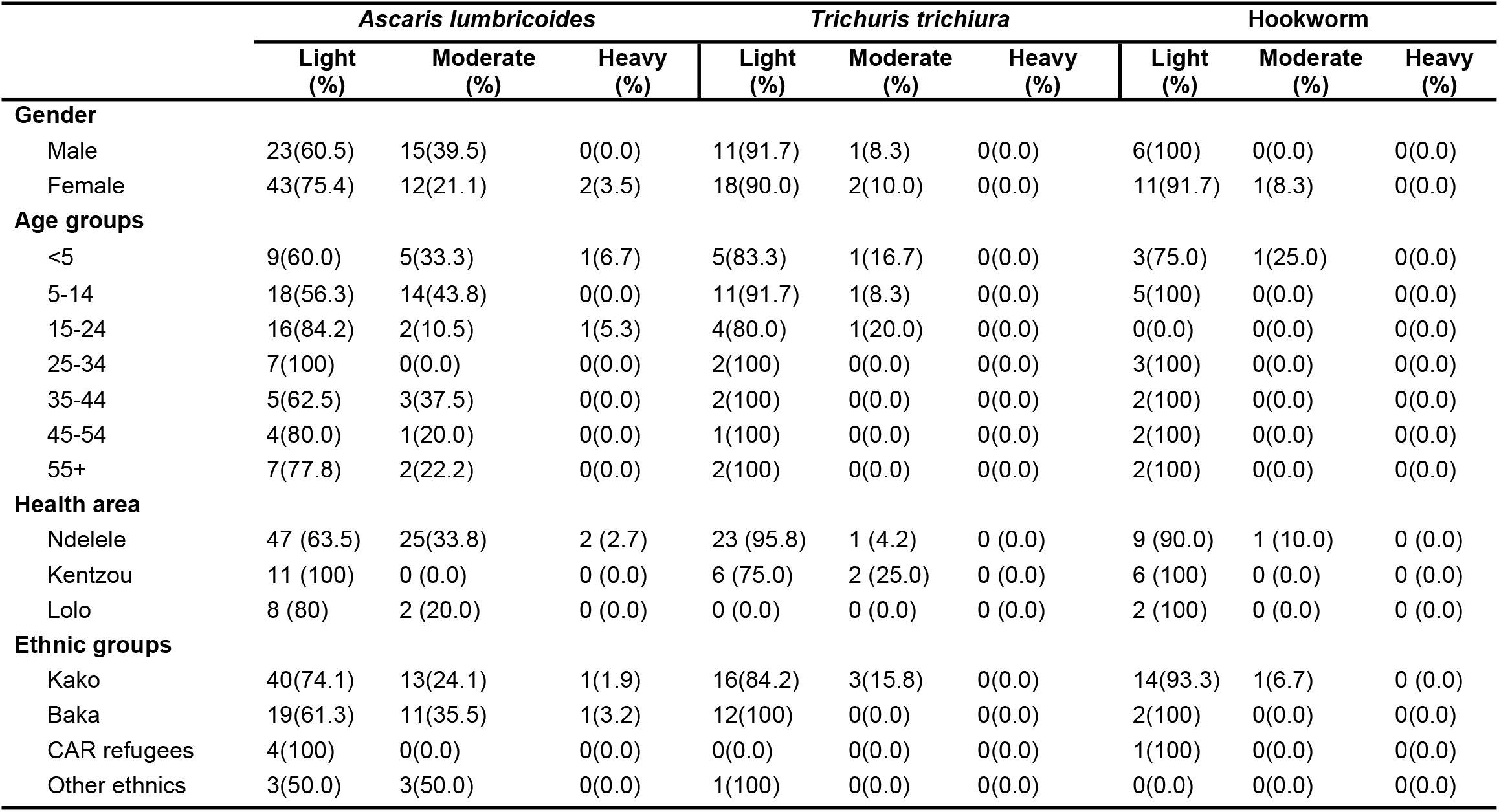
Intensity of helminth infections.

**Table 5.**
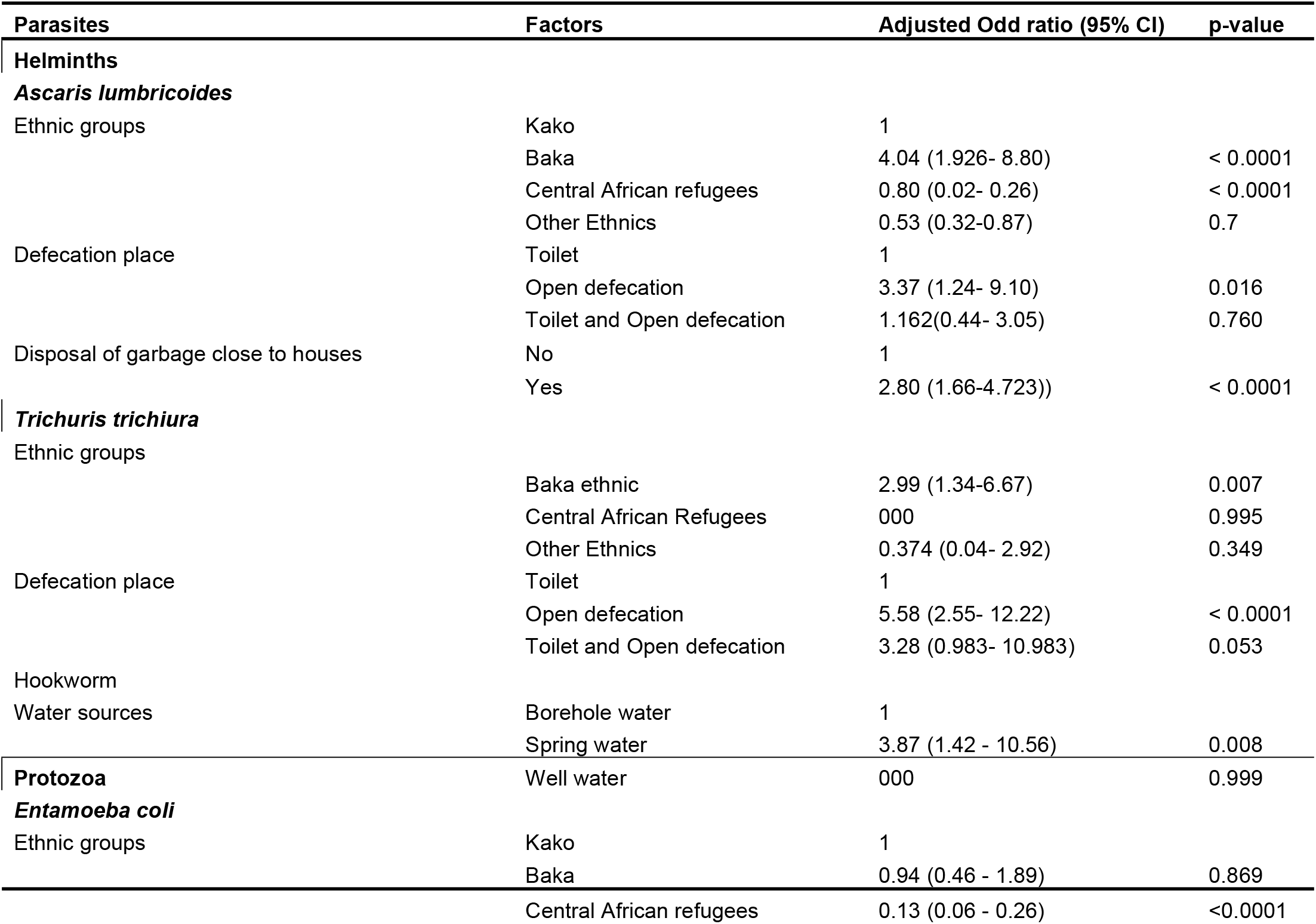
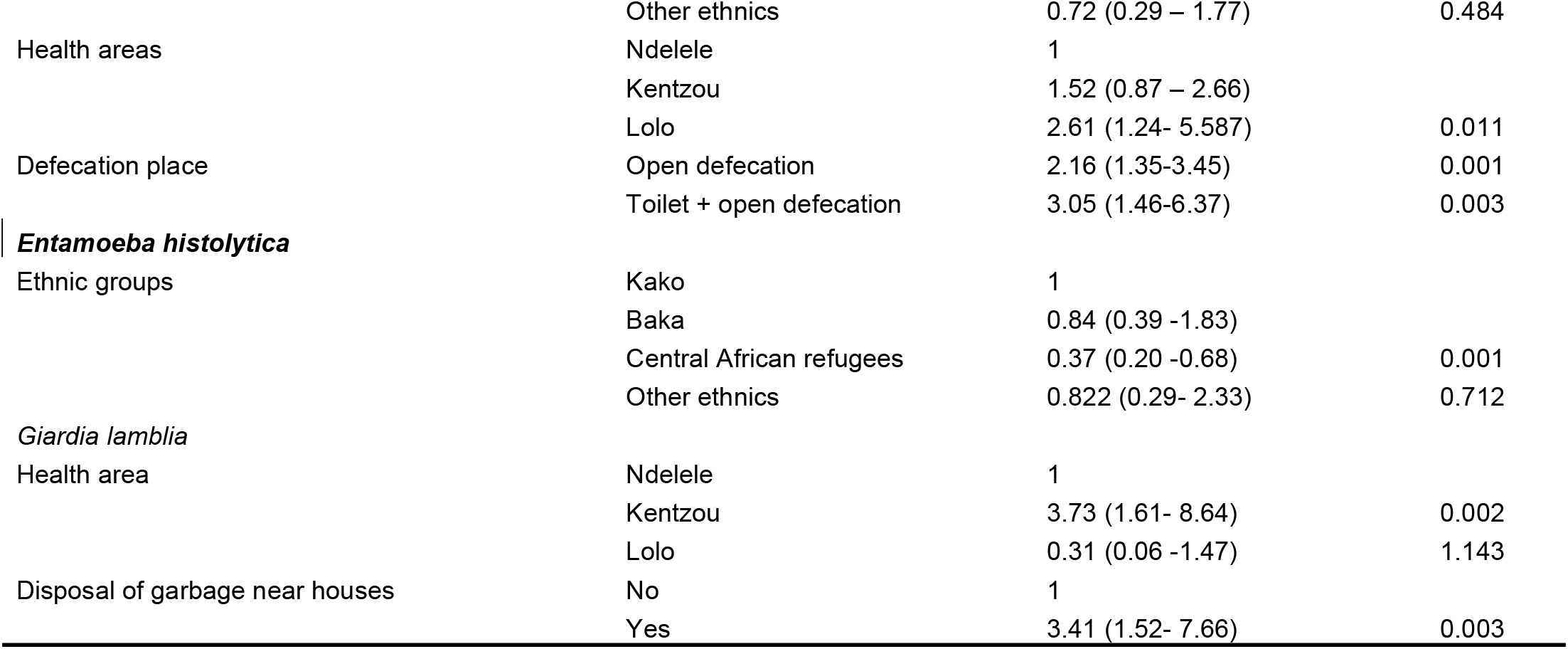
Risk factors Parasites.

## Discussion

The present study aimed to understand the epidemiology of intestinal parasitic infections in the Ndelele, Kentzou and Lolo Health Areas of the Ndelele Health District. Overall, 74.9% of participants were infected by at least one intestinal parasite species, which is similar to the infection rate (88.2%) in the neighboring areas of Central African Republic [24]. By contrast this prevalence was very high compared to those obtained by Mbuh *et al*. (31.0%) and Denecho *et al*. (59.1%) in the South-West Region of Cameroon [25-26] and Saoting *et al*. (30.0%) in the North Region of Cameroon [27]. This very high infection prevalence can be explained by the poor hygiene and sanitation, and reduced access to appropriate treatments in these landlocked areas. Indeed, a KAP survey conducted as part of this study revealed poor knowledge of interviewees about parasitic infection transmission. Those who identified food as the main mode of contamination were making reference to the consumption of sweet fruits (68.1%) such as mango or banana, contaminated food (12.9%) or water (10.7%). In addition, the KAP survey also revealed poor practice of hand hygiene as 96.6% of enrollees was practicing hand washing without soap before each meal. Also, due to absence of latrines in most of the households (20.4%, an important proportion (32.7%) of the population used to practice open defecation that can contribute to the persistence of transmission. It is also worth to mention that most of the enrollees did not use appropriate treatment (drugs known to be effective) against intestinal parasitic infections, likely because of the difficulty to access to modern medicines, and when treatment campaigns are organized, they are only restricted to at-risk groups. Indeed, the use of traditional medicine can represent a viable option but the fact that they have limited knowledge of clinical signs of different infections can jeopardize the efficacy of traditional medicines.

Likewise, infection by single parasite species, polyparasitism was also important (57.2%) in the framework of this study conducted in rural communities compared to relatively low rates (2.1%-14.7%) of polyparasitism found in urban areas such as in Douala, the Cameroon economic city capital [28-29]. These observations confirm the results of previous studies demonstrating that polyparasitism is more important in rural areas compared to urban settings [28, 30, 31], likely because of poor adherence to WASH. Indeed, a positive association was is often reported between poor practice of hand washing and polyparasitism;, showing that people who never wash their hands with soap before each meal are about 12 times more at risk of intestinal parasitic infections than their counterparts who always wash their hands before meals.

As part of the present study, two vulnerable/special populations were enrolled, namely the Baka (Pygmies) and the Central African Refugees. Baka’s are qualified as “indigenous people” because of their culture which is very different of that of the “dominant” ethnic group (Kako) [32]. Although these two groups are vulnerable, their rate of intestinal parasitic infection were statistically different, the Baka being heavily infested (93.3% vs 53.5%), and highly polyparasitized (66.7% vs 29.9%). Unlike refugees, Baka (Pygmies) live in the forest in extreme poverty conditions. Indeed, the Baka ethnic group exhibited the worst results for WASH indicators while most of their households (87.5%) do not have latrines. As consequence 88.9% of Baka participant practiced open defecation known as a facilitating factor for the dissemination of parasite stages (eggs and cysts) responsible of human infections. This situation is exacerbated by the fact that most may not wash their hands before eating, although they have declared to do so. Previous studies conducted among “indigenous communities” demonstrated that they are most often affected by helminth infections such as *A. lumbricoides, T. trichiura* and hookworms [33, 34, 35] as it was found in this study. Contrarily to Baka, Central African refugees found in the Ndelele Health District are, for the vast majority were placed under the protection of the High Commissioner for Refugees benefiting from many facilities from Non-Governmental Organizations, including UNICEF, PRO-ACT Project, *Premiere Urgence Internationale*, Care, Red Cross, *Solidarités Internationales*. Central African refugees therefore benefit from donations such as latrines and boreholes construction [36-37], permanent education on environmental and personal hygiene to improve their living conditions, and appropriate treatments. This can explain the fact that the infection rates of the targeted parasitic infections are quite low among this special population.

## Conclusion

The present study provides useful data on infection rates and risk factors associated with intestinal parasitic infection in the Ndelele Health District. The results highlighted exceptionally high rates of infections in this landlocked. Lack of sanitation and poor hygiene can largely contribute to the endemicity of intestinal parasitic infections, especially among indigenous and hard-to-reach populations (Pygmies). Targeted and complementary control strategies appear compulsory to reach these populations and offer appropriate care to interrupt or at least reduce the transmission of these debilitating diseases.

## Data Availability

All data produced in the present work are contained in the manuscript

## Acknowledgements

The authors are thankful to all the population of the Ndelele Health District for their hospitality and their participation. We are grateful to all those who contributed in the realization of this study.

